# Uptake of minimal intervention dentistry among Romanian dental professionals and trainees: an exploratory cluster and network analysis

**DOI:** 10.64898/2026.06.20.26356122

**Authors:** Diana Paula Simionescu, Ruxandra Sfeatcu, Arina Vinereanu

## Abstract

**Background:** Minimal intervention dentistry (MID) is promoted as a prevention-oriented approach to caries management, but its integration into routine practice remains uneven. Existing research often examines MID-related knowledge, attitudes, or practices separately, offering limited insight into how these dimensions co-occur within individuals or are conditionally associated.

**Methods:** This exploratory cross-sectional survey examined multidimensional MID uptake among 327 Romanian dental students, residents, and specialists from five university centers. Ten MID-related scores were analyzed, including nine formative composites and one single-item peer-norm indicator. K-means clustering examined uptake profiles, and Gaussian graphical model network analysis with stepwise BIC selection examined conditional associations among constructs.

**Results:** A two-cluster solution was highly reproducible but modestly separated (*n* = 144 vs *n* = 183; average silhouette width = 0.13; mean Jaccard similarities = 0.92 and 0.94). The profiles reflected broadly lower versus higher uptake across knowledge-, belief-, and practice-related dimensions, while perceived peer norms for hygiene instruction showed the opposite pattern. Profile membership was not clearly patterned by gender, age band, professional status, or clinical experience. The primary network included 14 non-zero edges out of 36 possible edges, all positive; the strongest partial association linked diagnostic knowledge to diagnostic methods used in practice (*partial r* = .22). Familiarity, diagnostic knowledge, and general practices occupied more interconnected positions descriptively, but limited centrality stability precluded interpreting them as intervention targets.

**Conclusions:** MID uptake in this sample was better represented as a continuum of modestly differentiated profiles than as sharply separated participant types. The findings provide an exploratory map of multidimensional MID uptake and may inform future survey validation, implementation research, and dental education studies. Because the study was cross-sectional, convenience-sampled, and based on self-report, findings should be interpreted as hypothesis-generating rather than causal or population-representative.

## Introduction

In contemporary dental practice, advances in cariology and restorative materials have supported a shift toward non-invasive and minimally invasive approaches that prioritize caries control and the preservation of healthy tooth structure [1]. The available evidence nevertheless needs to be interpreted in relation to specific clinical indications. For example, in primary teeth with dentinal caries, selected minimally invasive approaches have been shown to support caries arrest and clinical management when compared with no treatment or conventional restorative care [2]. Given the continuing global burden of dental caries [3] and persistent oral health inequalities [4], prevention-oriented care remains internationally relevant to dental practice and education.

Assessing dental professionals’ knowledge, attitudes, and practices regarding MID is therefore essential for identifying gaps between evidence and clinical application and for informing curriculum development. Although attitudes toward MID are generally positive, prior research indicates deficiencies in understanding and applying specific techniques [5–9]. Educational strategies that combine structured hands-on training with attention to the professional contexts in which MID is learned and practiced may therefore be important for supporting its routine integration in dental care.

These educational priorities are consistent with broader public-health and workforce-oriented directions in oral health, including prevention, oral-health promotion, evidence-informed decision-making, and competency-based professional training [10, 11]. For dental education, this suggests that MID is better framed not only as a set of isolated techniques, but as an integrated approach linking diagnostic reasoning, caries-risk assessment, preventive knowledge, minimally invasive clinical decision-making, and patient-centered care [12].

Existing research typically examines MID-related knowledge, attitudes, or practices separately and often provides a predominantly descriptive view [5, 6, 9, 13–15]. This limits understanding of how different dimensions of MID uptake co-occur within individuals and how they relate to one another as part of an educationally relevant configuration. It therefore remains unclear whether interpretable profiles of MID uptake can be identified among dental professionals and trainees, and which MID dimensions are most closely connected within this broader pattern. Addressing this gap may help educators move beyond one-size-fits-all training toward more targeted, context-sensitive educational strategies.

Accordingly, the aims of this exploratory study were to (1) examine whether interpretable profiles of MID uptake could be identified among dental professionals and trainees based on multiple MID-related constructs, and (2) examine the interrelationships among these constructs to better understand their configuration within dental education and practice. To this end, we conducted a cross-sectional survey using complementary profile-centered and network-analytic approaches during the early implementation period of the *Pain-Free Dental Treatment (PaFein+) Teaching Model and Its Effect on Dental Treatment of Children*, an Erasmus+ Cooperation Partnerships project that integrates minimally invasive dentistry to improve children’s experience of dental care.

## Methods

### Participants and Sampling

The study targeted dental professionals and trainees in Romania. Eligibility criteria included being 18 years of age or older, having current clinical activity or being enrolled in dental training, and providing informed consent for participation in the study. We used convenience sampling. The study was conducted within an international educational project with English as the working language. Invitations were disseminated electronically to residents in pediatric dentistry and other dental specialties, final-year dental students, and specialists involved in academic teaching from five university centers in Romania (Bucharest, Cluj-Napoca, Iași, Târgu Mureș, and Timișoara). The final dataset did not retain respondent-level indicators of recruitment source or university center; therefore, direct evaluation of source-specific or center-specific effects was not possible. Because participation was anonymous and the link circulated without a sampling frame, a response rate could not be estimated.

The final sample included 327 participants (83.2% women): 59.3% students, 22.9% residents, and 17.7% specialists. Most were aged 20–30 years (77.1%), with 12.2% aged 31–40 and 10.7% aged ≥41; 22.3% reported 5 or more years of clinical practice. Regarding specialty, 196 participants (59.9%) had no specialty recorded, almost all of whom were students. Among practitioners (residents/specialists; *n* = 133), 131 reported at least one specialty. Because two practitioners reported mixed specialties, specialty categories were coded as non-mutually exclusive. Pediatric dentistry/pedodontics was the most commonly reported specialty (*n* = 55, 41.4%), followed by general dentistry (*n* = 46, 34.6%), prosthodontics (*n* = 15, 11.3%), endodontics (*n* = 6, 4.5%), periodontology (*n* = 5, 3.8%), orthodontics/dentofacial orthopedics (*n* = 3, 2.3%), dentoalveolar surgery (*n* = 2, 1.5%), and esthetic dentistry (*n* = 1, 0.8%). Two practitioners did not report a specialty.

No a priori sample-size calculation was conducted because the study was exploratory and recruitment was constrained by the available project-related educational and professional networks; all complete eligible responses were included.

### Study Design

We employed a cross-sectional survey design. The questionnaire was distributed online during two recruitment periods in 2024: an early period from March to May and a later period from October to November. Distribution occurred through routine educational and professional channels, and responses were completed online. Timestamp patterns indicated a mixture of clustered completion windows and more dispersed asynchronous responses. In view of the convenience-based recruitment and limited respondent-level contextual metadata, the findings should be interpreted as exploratory.

### Survey Instrument and Variables

#### Instrument development

The questionnaire was developed in English and refined through iterative expert review within the PaFein+ consortium. For the present Romanian administration, the questionnaire was translated into Romanian by consortium members with experience in professional dental terminology and bilingual academic materials, then refined iteratively to preserve the meaning of the English items and ensure clarity in Romanian. A formal forward–back translation procedure or independent linguistic validation was not conducted.

Items assessed awareness, training, knowledge, beliefs, practices, and perceived peer norms related to MID using closed-ended response formats. MID constructs were operationalized as formative composite scores based on theoretical and clinical rationale. The items within each composite were treated as complementary indicators of the relevant domain, such as training exposure, diagnostic knowledge, or practice behaviors, rather than as interchangeable manifestations of a single latent trait. Therefore, component quality was evaluated through distribution checks, floor/ceiling inspection, collinearity diagnostics, leave-one-out sensitivity analyses, and item–index directionality checks. Across the multi-item composites, maximum component-level VIF values ranged from 1.09 to 2.81, indicating no problematic collinearity, and no component showed a negative item–index correlation (Table S1 in *S1 Appendix*).

#### MID Variables

Items were directionally aligned so that higher values indicate greater endorsement/use. Items phrased in opposition to MID principles were reverse-coded, and composite scores were computed as the mean of constituent items after domain-specific coding and, where needed, linear rescaling to a common metric. The complete item wording (in English and Romanian) and response coding are provided in Table S2 in *S1 Appendix*.

*Yes/No/Not sure* responses were coded according to item function. For *familiarity* and *knowledge* items, “Not sure” was treated as an intermediate response because uncertainty may indicate partial awareness rather than definite absence of knowledge. For practice items, “Not sure” was treated as non-endorsement, because uncertainty did not provide evidence that the behavior was performed. Similarly, for efficacy items, “Don’t know/Not familiar” was coded as non-endorsement of effectiveness.

For composites combining items with different response ranges, scores were linearly rescaled to a shared theoretical metric before averaging. Specifically, 0–4 Likert items included in composites otherwise scored on a 0–2 metric were divided by 2. This transformation is equivalent to min–max rescaling from 0–4 to 0–2 using the theoretical scale endpoints, while avoiding sample-dependent normalization based on observed minimum and maximum values. Conversely, 0–2 items included in the *attitudes* composite were multiplied by 2 to retain that composite on a 0–4 metric. These coding and rescaling decisions were specified before the main analyses and are fully documented in Table S2 in *S1 Appendix*.

##### Familiarity with MID

A formative composite reflecting self-reported familiarity with MID principles and standard-of-care status; higher scores indicate greater familiarity.

##### MID Training

A formative composite capturing exposure during dental education to MID-related preventive and minimally invasive techniques; higher scores indicate broader training exposure.

##### Diagnosis Knowledge

A formative composite assessing awareness of caries diagnostic technologies consistent with MID; higher scores reflect stronger diagnostic knowledge.

##### Diagnostic Methods (Own Practice)

A formative composite reflecting current diagnostic behaviors, with higher scores indicating greater alignment with MID-consistent practice.

##### Caries Classification Systems (Own Practice)

A formative composite capturing the use of caries classification systems in clinical practice, with higher scores indicating more contemporary approaches.

##### General Practices (Own Practice)

A formative composite assessing engagement in preventive and risk-based clinical practices; higher scores reflect broader adoption of MID-consistent practices.

##### Attitudes toward MID

A formative composite assessing personal views toward MID principles, with higher scores indicating more positive attitudes toward MID integration.

##### Efficacy Beliefs

A formative composite reflecting perceived efficacy of preventive, non-restorative, and minimally invasive approaches, both general and tooth-specific (deciduous and permanent teeth); higher scores indicate stronger efficacy beliefs.

##### Preventive Knowledge

A formative composite of two items assessing core preventive principles; higher scores indicate stronger preventive knowledge.

##### Peer Norms (Hygiene Instruction)

A single-item, reverse-scored measure capturing perceived peer endorsement of hygiene instruction as a caries-risk management strategy; higher scores indicate stronger perceived endorsement.

## Data Analysis

We used two complementary exploratory approaches. First, k-means clustering was employed as a profile-centered method to group participants with similar response patterns across the MID scores. In practical terms, this analysis asked whether participants tended to form interpretable patterns of MID uptake across several dimensions considered together, rather than examining each dimension separately. Second, network analysis was applied to examine the structure of conditional relationships among MID composites—an approach that combines multivariate statistics and network science to represent dependency patterns among variables [16]. Thus, clustering described how MID-related dimensions were configured within participants, whereas network analysis described how the constructs were interconnected across the sample.

Before implementing cluster and network analyses, all MID score variables were standardized (z-scores) to avoid undue influence from scale differences. All analyses were conducted in R (version 4.4.1) and RStudio (version 2024.12.0+467). A list of the main statistical software packages used, along with their references, is provided in Table S3 in *S1 Appendix*.

### Cluster analysis

We applied k-means clustering to the 10 z-standardized MID scores using the Hartigan–Wong algorithm [17] with Euclidean distance, 1,000 random starts, and a maximum of 1,000 iterations per start. K-means clustering assigns participants to groups so that participants within the same group have more similar response patterns to one another than to participants in other groups. To determine the number of clusters, we evaluated candidate solutions with 2–15 clusters using three internal criteria: (a) the elbow plot of total within-cluster sum of squares, (b) the average silhouette width on the Euclidean distance matrix, and (c) a majority-vote panel of indices.

After selecting the number of clusters, we refitted the final model and described profiles by standardized centers. These centers represent the average standardized score of each cluster on each MID score and were used to interpret the substantive meaning of the profiles. Cluster stability was evaluated using a nonparametric bootstrap of 1,000 resamples. For each resample, we reran k-means with 100 random starts and reported the mean Jaccard similarity and the number of cluster dissolutions. Higher Jaccard values indicate that similar clusters are recovered across resampled datasets.

Between-profile differences across the 10 MID scores were tested using two-sided, two-sample Welch’s *t*-tests. *P*-values were adjusted for the 10 comparisons using the Benjamini–Hochberg procedure to control the false discovery rate (FDR) at 0.05 [18]. Effect sizes were reported as Hedges’ *g*. We further examined demographic balance (gender, age band, clinical experience band, and professional status) using χ² tests with Cramér’s *V* as the effect size. To satisfy the expected-count assumptions, the single “prefer not to say” gender response was treated as missing for that test.

To examine whether the findings were sensitive to recruitment period and respondent composition, we conducted sensitivity analyses based on two recruitment periods derived from response timestamps (March‒May 2024 vs October‒November 2024). We first compared respondent composition across periods using χ² tests with Cramér’s *V*. We then compared MID variables across periods using Welch’s *t*-tests with Benjamini–Hochberg correction and Hedges’ *g*, and fitted linear models estimating recruitment-period differences after adjustment for professional status. Cluster membership was compared across recruitment periods using a χ² test with Cramér’s *V*. As an additional sensitivity check, we used logistic regression to estimate the association between recruitment period and binary cluster membership, both before and after adjustment for professional status.

### Network analysis

We estimated a Gaussian Graphical Model (GGM) among the MID composites directly linked to individual knowledge, training, beliefs, and practice, with *peer norms for hygiene instruction* excluded *a priori* because it was conceptualized as a single-item contextual indicator rather than a multi-item MID competence domain. In this network, *nodes* represent MID composites, and *edges* represent partial associations between pairs of nodes after accounting for all other nodes. To assess whether excluding this node materially altered the overall network structure, we also re-estimated the network in a sensitivity analysis that included peer norms; these results are reported in *S1 Appendix* (Figs S1‒S2).

The estimator was the stepwise Bayesian Information Criterion (BIC). As described in [19], the algorithm: (i) uses graphical LASSO, a penalty method [20], to generate a sequence of regularized candidate networks (by default, 100 models ranging from sparse to dense); (ii) re-estimates each candidate without regularization, constraining absent edges to zero, and selects the best model by BIC; (iii) performs stepwise edge additions or removals—each time re-estimating the unregularized model—until no further BIC improvement is possible.

Stepwise BIC was selected based on simulation evidence indicating higher specificity at moderate sample sizes [19, 21]. Rank-based Spearman correlations were used to accommodate non-normality and ordinal measurement. For descriptive purposes, node strength, closeness, and betweenness were computed, but their interpretation was guided by stability analyses [21]. Network accuracy and centrality stability were evaluated using standard nonparametric bootstrapping procedures [22].

## Ethical Considerations

The study was approved by the Research Ethics Committee of Carol Davila University of Medicine and Pharmacy, Bucharest, Romania, approval no. 11580/30.04.2024. All procedures were conducted in accordance with the Declaration of Helsinki, the EU GDPR (2016/679), and applicable national regulations. An information page preceding the survey explained to participants the study’s purpose, the assurance of confidentiality, and that their responses would be used solely for aggregate statistical analyses in scientific research. Consent was provided by continuing to the next page and by submitting the survey. Participation was voluntary, and participants could discontinue at any time before submission without penalty. No incentives were offered. The analysis dataset contained no patient-level data and no direct personal identifiers. Optional contact details provided by participants for follow-up purposes were stored separately and were not included in the analysis dataset. For students and residents, non-participation had no academic or supervisory consequences.

## Results

### Participant profiles based on MID uptake

Model selection supported a cautious two-cluster solution. A majority-rule panel of internal indices favored two clusters (seven indices), with comparable support for nine and scattered support for other solutions. The average silhouette width also peaked for two clusters, but its value was low (0.13), and the elbow plot showed no pronounced knee. We therefore retained *k* = 2 as the most parsimonious and interpretable exploratory solution, while recognizing that the low silhouette indicated modest separation rather than sharply distinct participant types.

The final clusters contained 144 and 183 participants. Consistent with the low silhouette value, separation was modest (mean silhouettes 0.08 and 0.18; the partition captured approximately 14% of the total variation across MID scores). However, the solution was highly reproducible under nonparametric bootstrap resampling (1,000 resamples): mean Jaccard similarities were 0.92 (Cluster 1) and 0.94 (Cluster 2) with no dissolutions. Thus, the solution should be interpreted as a reproducible, broad profile pattern consistent with a continuum of MID uptake, rather than as evidence of strongly separated subgroups.

Standardized centers (Fig 1) identified two interpretable profiles. Cluster 1 showed lower values across the MID spectrum—less *familiarity* and *training*, lower *knowledge* (general and diagnostic-specific), fewer MID-consistent *practices* (general and diagnostic-specific), and more reserved *attitudes* and *efficacy beliefs*—yet reported higher perceived *peer norms for hygiene instruction*. Cluster 2 showed the opposite pattern: broadly higher MID uptake on those dimensions, alongside lower perceived *peer norms for hygiene instruction*.

**Fig 1.**
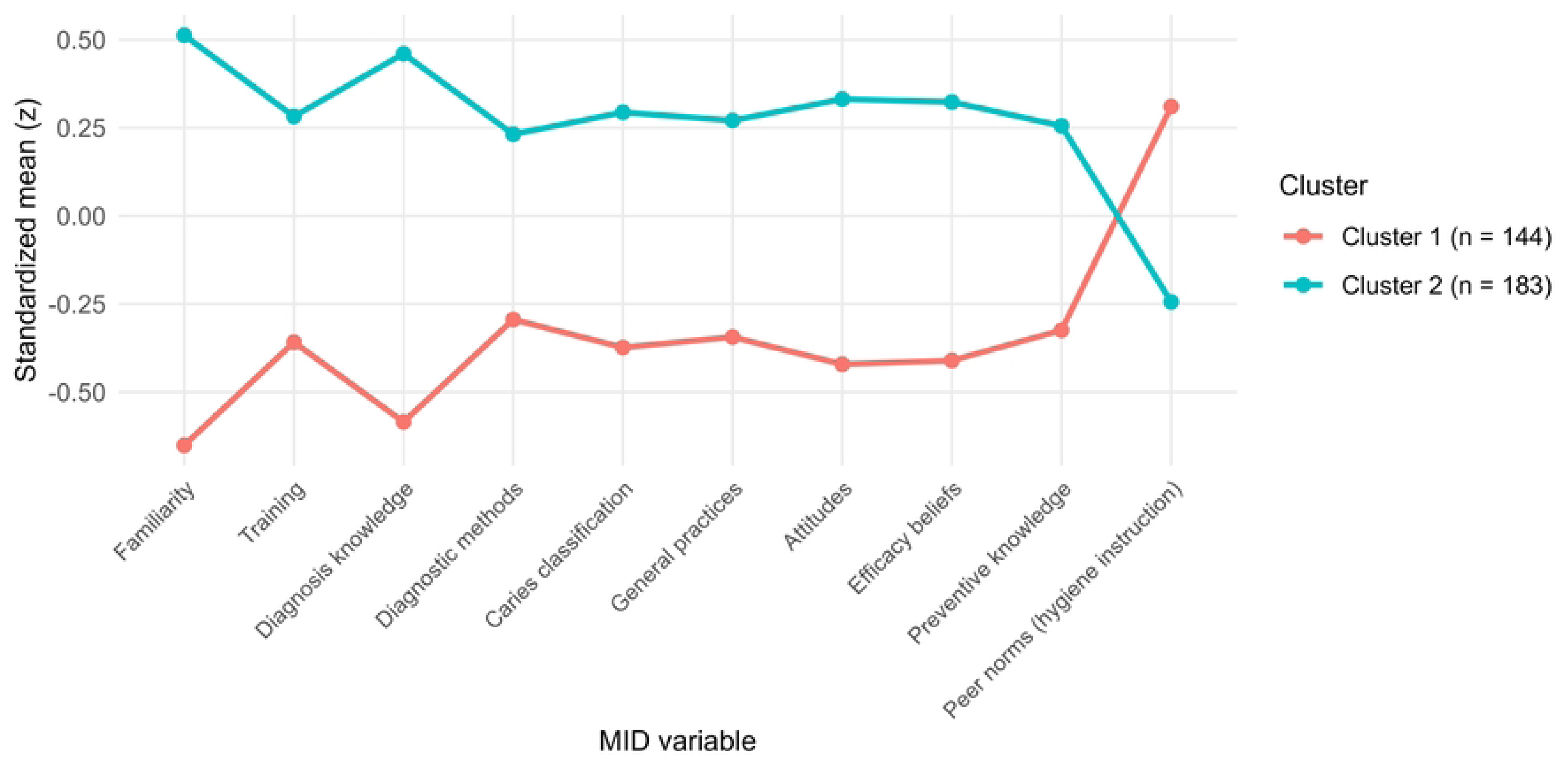
Cluster profiles across MID variables. Higher values indicate greater endorsement/use on each MID score. Points are standardized cluster centers (z-scores). *N* = 327.

Across the 10 scores, clusters differed significantly on every measure after Benjamini–Hochberg adjustment (all BH-adjusted *p*-values < .001), with medium-to-large effects (Table 1). The largest gaps were observed for *MID familiarity* (*|g|* = 1.42) and *diagnostic knowledge* (*|g|* = 1.22), whereas the smallest differences were found for *diagnostic methods used in practice* (*|g|* = 0.54) and *peer norms for hygiene instruction* (*|g|* = 0.57).

**Table 1.** Standardized centroids by cluster and between-profile differences on the 10 MID scores.

| MID variables | Cluster 1<br>($n = 144$) | | Cluster 2<br>($n = 183$) | | Welch's two-sample $t$-tests | | | |
| --- | --- | --- | --- | --- | --- | --- | --- | --- |
| | $M$ | $SD$ | $M$ | $SD$ | $df$ | $t$ | Hedges' $g$ | $p$ (BH) |
| Familiarity | -0.65 | 0.87 | 0.51 | 0.77 | 287.2 | -12.6 | 1.42 | $< .001$ |
| Training | -0.36 | 0.88 | 0.28 | 1 | 320.5 | -6.15 | 0.67 | $< .001$ |
| Diagnosis knowledge | -0.58 | 0.83 | 0.46 | 0.88 | 314.0 | -11.04 | 1.22 | $< .001$ |
| Diagnostic methods | -0.29 | 1.12 | 0.23 | 0.83 | 254.8 | -4.71 | 0.54 | $< .001$ |
| Caries classification | -0.37 | 0.88 | 0.29 | 0.99 | 319.8 | -6.43 | 0.70 | $< .001$ |
| General practices | -0.34 | 1.22 | 0.27 | 0.68 | 210.7 | -5.43 | 0.64 | $< .001$ |
| Attitudes | -0.42 | 0.95 | 0.33 | 0.92 | 302.5 | -7.25 | 0.81 | $< .001$ |
| Efficacy beliefs | -0.41 | 0.99 | 0.32 | 0.88 | 287.8 | -6.96 | 0.79 | $< .001$ |
| Preventive knowledge | -0.32 | 1.09 | 0.26 | 0.84 | 261.3 | -5.26 | 0.60 | $< .001$ |
| Peer norms (HI) | 0.31 | 0.96 | -0.24 | 0.96 | 306.6 | 5.17 | -0.57 | $< .001$ |

*Note.* HI = hygiene instruction. Values are means and standard deviations of z-standardized scores. Welch's two-sample $t$ -tests (two-sided) are reported; $df$ values use the Satterthwaite approximation. Effect sizes are Hedges' $g$ ; positive values indicate higher scores in the higher-uptake profile (Cluster 2). $p$ (BH) = Benjamini–Hochberg FDR-adjusted $p$ across the 10 tests.

Table 2 shows the distribution of MID profiles by participant characteristics and recruitment period. Cluster membership did not differ by gender, age band, professional status, or clinical experience. In sensitivity analyses, restricting to practitioners (residents vs. specialists) again showed no association, χ²(1, *N* = 133) = 0.60, *p* = .44, Cramér’s *V* = .08; excluding the smallest experience band (under 5 years) yielded the same conclusion, χ²(2, *N* = 73) = 2.63, *p* = .27, Cramér’s *V* = .19.

**Table 2.** Distribution of MID profiles by participant characteristics, recruitment period, and χ² tests of association.

| Characteristic | Participant distribution |  | Chi-squared tests |  |  |  |
| --- | --- | --- | --- | --- | --- | --- |
| | Cluster 1<br>(*n* = 144) | Cluster 2<br>(*n* = 183) | $\chi^2$ | *df* | *p* | Cramér's *V* |
| Gender |  |  | 0.00 | 1 | .96 | .01 |
| Female | 120 (44.12%) | 152 (55.88%) |  |  |  |  |
| Male | 23 (42.59%) | 31 (57.41%) |  |  |  |  |
| Age |  |  | 2.59 | 2 | .27 | .09 |
| 20–30 years | 116 (46.03%) | 136 (53.97%) |  |  |  |  |
| 31–40 years | 13 (32.50%) | 27 (67.50%) |  |  |  |  |
| 40+ years | 15 (42.86%) | 20 (57.14%) |  |  |  |  |
| Professional status |  |  | 5.57 | 2 | .06 | .13 |
| Student | 95 (48.97%) | 99 (51.03%) |  |  |  |  |
| Resident | 25 (33.33%) | 50 (66.67%) |  |  |  |  |
| Specialist | 24 (41.38%) | 34 (58.62%) |  |  |  |  |
| Clinical experience |  |  | 4.39 | 3 | .22 | .12 |
| < 5 years | 117 (46.06%) | 137 (53.94%) |  |  |  |  |
| 5–10 years | 11 (37.93%) | 18 (62.07%) |  |  |  |  |
| 11–15 years | 3 (20.00%) | 12 (80.00%) |  |  |  |  |
| > 15 years | 13 (44.83%) | 16 (55.17%) |  |  |  |  |
| Recruitment period |  |  | 8.75 | 1 | .003 | .17 |
| March–May | 58 (35.58%) | 105 (64.42%) |  |  |  |  |
| October–November | 86 (52.44%) | 78 (47.56%) |  |  |  |  |

*Note.* Percentages are row percentages within each participant-characteristic or recruitment-period category. For 2×2 tables, *p*-values are computed using Yates' continuity correction. Cramér's *V* effect sizes are computed from the uncorrected Pearson $\chi^2$ . One "prefer not to say" gender response was excluded. The age bands 41–50 and 50+ were collapsed into 41+. Recruitment periods were derived from response timestamps. *N* = 327.

Recruitment-period sensitivity analyses indicated that the two recruitment periods were not compositionally equivalent (Tables S4–S5 in *S1 Appendix*). Compared with the early period, the later period included a higher proportion of students (70.7% vs 47.9%) and lower proportions of residents and specialists, χ²(2, *N* = 327) = 17.73, BH-adjusted *p* < .001, Cramér’s *V* = .23. Age and clinical experience also differed by recruitment period, whereas gender did not. Several MID composites were lower in the later period, particularly *general practices*, *caries classification systems*, *attitudes toward MID*, and *preventive knowledge*; these differences remained after adjustment for professional status (standardized β range: -0.43 to - 0.30, BH-adjusted *p*-values = .001–.020). Cluster membership also differed by recruitment period, χ²(1, *N* = 327) = 8.75, *p* = .003, Cramér’s *V* = .17, with a higher proportion of lower-uptake participants in the later period (52.4% vs 35.6%). In a logistic model predicting membership in the higher-uptake cluster, this association persisted after adjustment for professional status, *OR* = 0.54, 95% *CI* [0.34, 0.85], *p* = .008. These findings indicate recruitment-period heterogeneity in this convenience sample. Because respondent-level recruitment source and university center were not recorded, the observed differences cannot be attributed to a specific recruitment channel, institutional context, or causal effect of calendar time.

### Interconnections among MID constructs

The partial-correlation network (GGM) among the nine education-related MID composites was moderately sparse: 14 out of 36 edges were non-zero (all positive; mean |edge| = .06; Fig 2). The numerically strongest edge linked *diagnostic knowledge* with *diagnostic methods* (*partial r* = .22). Two additional edges were of similar magnitude (*familiarity* with *efficacy beliefs* and *familiarity* with *caries classification*, both *r* = .19). The remaining non-zero edges formed a comparable band (*partial r* = .15–.17). Pairwise edge-difference tests indicated that none of the edges were reliably distinguishable from the others.

**Fig 2.**
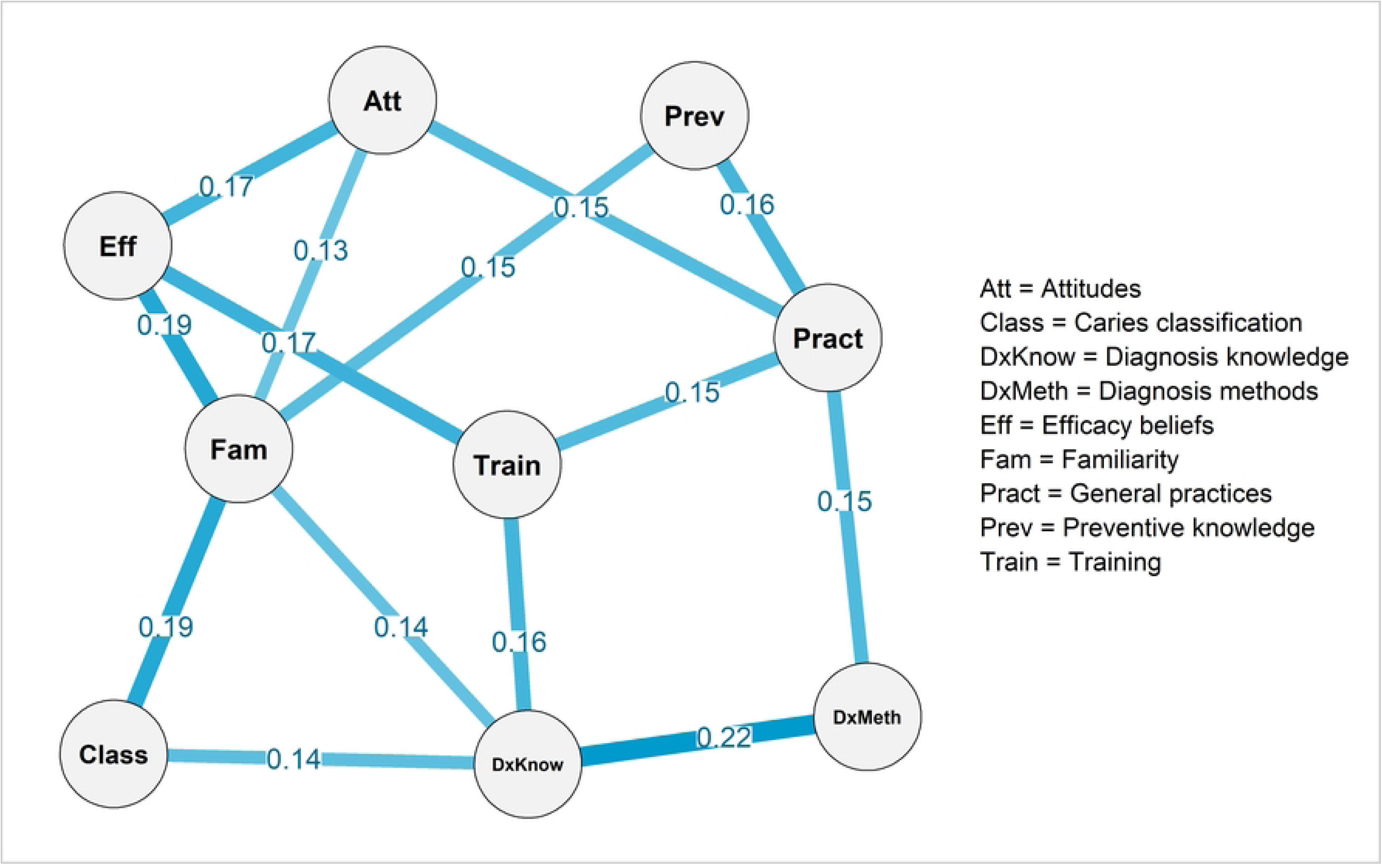
Partial-correlation network among MID constructs. The network was estimated using stepwise BIC model selection. Nodes are standardized composite scores. Edges are partial correlations (rank-based; Spearman input), conditioned on all other variables. Edge thickness reflects absolute magnitude. All edges were positive. *N* = 327.

Edge-weight accuracy analyses based on nonparametric bootstrapping (95% confidence intervals; Fig 3A) showed that non-zero edges identified by the stepwise BIC model generally exhibited bootstrap means closely aligned with their corresponding sample estimates. Several edges estimated as zero in the original network displayed small non-zero bootstrap means, indicating sensitivity to sampling variability.

**Fig 3.**
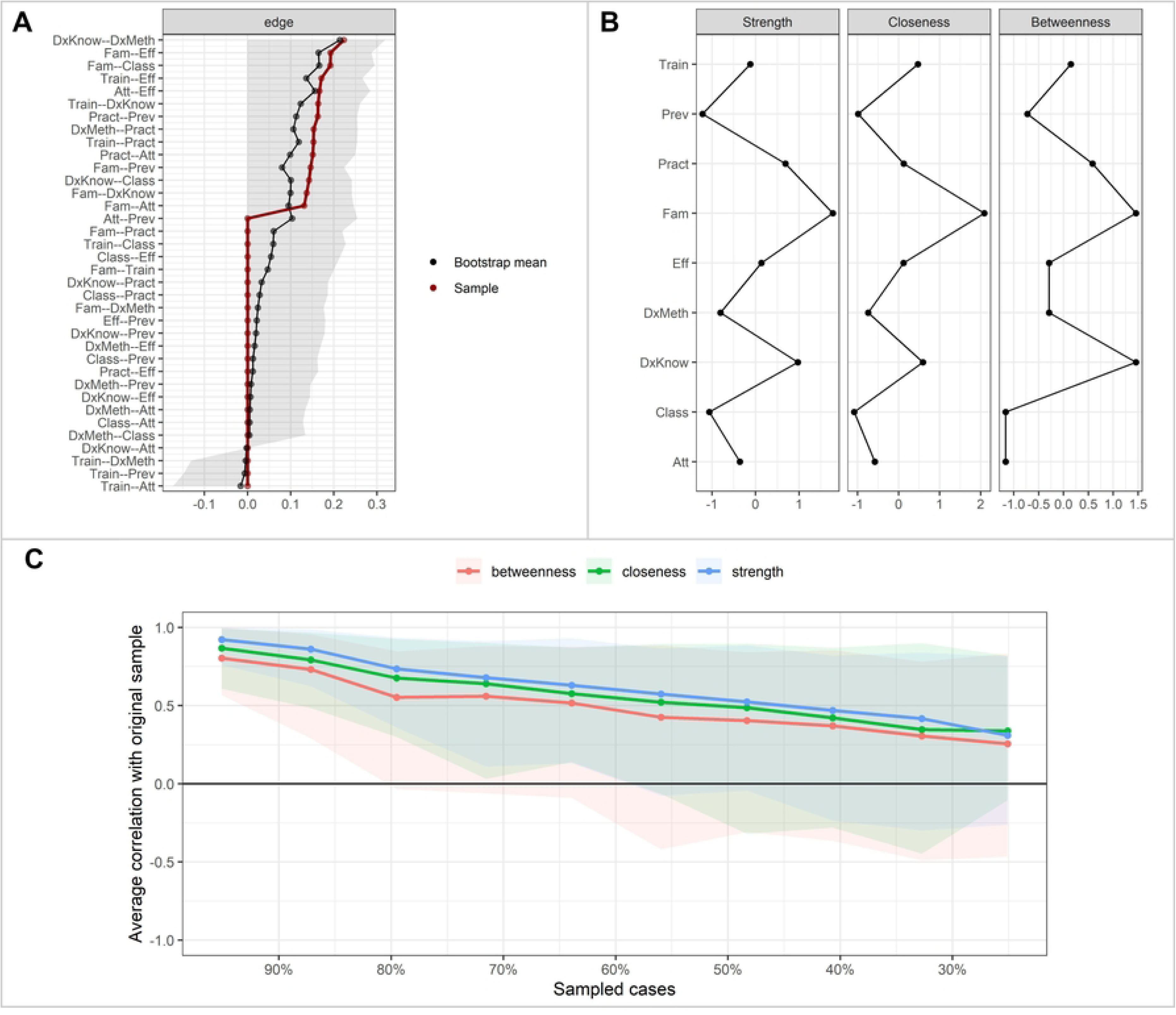
Network accuracy, centrality, and stability analyses. *Panel A* depicts edge-weight accuracy based on nonparametric bootstrapping (1,000 samples), showing original sample estimates (red), bootstrap means (black), and 95% confidence intervals, with edges ordered by their estimated weight. *Panel B* shows standardized centrality indices (z-scores) for strength, closeness, and betweenness across network nodes. *Panel C* presents case-dropping bootstrap results for centrality stability, plotting the average correlation between subset-sample and full-sample centrality estimates as a function of the proportion of cases retained; shaded areas indicate variability across bootstrap samples.

Standardized centrality indices (Fig 3B) indicated that *familiarity* had the highest strength, followed by *diagnostic knowledge* and *general practices*. *Efficacy beliefs*, *training*, and *attitudes* occupied intermediate positions, whereas *diagnostic methods*, *caries classification*, and *preventive knowledge* showed comparatively low strength. *Familiarity* also ranked highest in terms of closeness and betweenness.

However, case-dropping bootstrap analyses revealed limited centrality stability (Fig 3C). At the conventional threshold of *r* = .70, Correlation-Stability coefficients (CS) were approximately 0.05 for both closeness and strength, and 0 for betweenness. Stability improved only modestly at lower thresholds: at *r* = .50, CS values reached 0.20 for strength, 0.13 for closeness, and 0.05 for betweenness, while at *r* = .30, strength increased to 0.36, with closeness at 0.20 and betweenness remaining at 0.05. As none of the centrality indices met recommended stability benchmarks, all centrality findings (including strength) should be interpreted descriptively.

In a sensitivity network that included perceived peer norms for hygiene instruction (Figs S1–S2 in *S1 Appendix*), the overall structure remained largely unchanged: the strongest edges and the descriptive prominence of familiarity, diagnostic knowledge, and general practices were similar to those in the primary network. Peer norms entered the network as a peripheral node with a single negative edge to familiarity (*partial r* = -0.14), while centrality estimates remained insufficiently stable for substantive interpretation.

## Discussion

Contemporary oral healthcare faces a three-pronged imperative: reduce the disease burden at the population level [3]; deliver restorative treatment that is biologically respectful, prevention-oriented, and feasible in everyday clinical settings [2]; and attenuate oral health inequalities [4]. In this context, MID, widely promoted as a paradigm shift integrating prevention, early detection, risk-based decision-making, and patient-centered care [7], appears relevant.

However, MID is not a single skill. It is a constellation of interdependent competencies and beliefs, and despite broad professional endorsement, the uptake of its principles in practice remains uneven and challenging [2, 8, 23]. Dental education and continuing professional development are central to this challenge. Identifying less-developed MID dimensions, their interrelations, and the groups or contexts in which they appear may help educators design training that is better aligned with learners’ needs rather than relying on generic approaches.

This study examined how MID is understood, endorsed, and reported in practice among a sample of Romanian dental professionals and trainees, using a profile-centered clustering approach complemented by a network analysis of MID constructs. Moving beyond prior research that typically assessed MID-related knowledge, attitudes, or practices in isolation, our findings provide an integrated view of how such dimensions co-occur and are conditionally associated within a broader competence configuration relevant to dental education and training.

### Profiles of MID uptake

The cluster analysis revealed two reproducible and interpretable but modestly separated profiles of MID uptake. One group showed lower familiarity, training exposure, knowledge, efficacy beliefs, and self-reported practices, whereas the other displayed higher endorsement across these domains. The profiles were not associated with gender, age band, professional status, or years of clinical experience in the main demographic comparisons. However, recruitment-period sensitivity analyses showed that profile membership varied across the two recruitment periods, which also differed in respondent composition. Taken together, these findings suggest that the distribution of profiles may be sensitive to recruitment context and sample composition, rather than being clearly patterned by demographic or seniority indicators. The profiles should therefore be interpreted as exploratory patterns rather than stable population subgroups. From an educational perspective, this is consistent with broad curriculum strategies that address MID uptake across learner and practitioner groups, while remaining attentive to local training contexts and individual learning needs.

An interesting and somewhat counterintuitive finding was that participants in the lower-uptake cluster reported stronger perceived peer norms for hygiene instruction than those in the higher-uptake cluster. One cautious interpretation is that basic preventive advice may be more visible and socially reinforced across dental settings than more complex MID-related practices. This offers one possible explanation for why lower-uptake participants perceived stronger peer endorsement of hygiene instruction. Conversely, participants with higher MID uptake may evaluate peer practices against more stringent standards for evidence-based hygiene instruction. Because the survey did not directly assess how respondents interpreted peer norms, this explanation should be treated as hypothesis-generating.

From a dental education perspective, this finding underscores the importance of moving beyond symbolic endorsement of prevention toward explicit teaching, modeling, and assessment of hygiene instruction quality. Future educational research could examine whether descriptive-norm feedback—that is, information about what trained peers actually do—and carefully framed messages about peers’ practices support the translation of preventive principles into clinical practice and strengthen professional norms around prevention.

Nevertheless, the two participant clusters were modestly differentiated, which appears more consistent with a gradient perspective of MID uptake than with the assumption of sharply separated groups among Romanian professionals and trainees. In other words, the findings point to a continuum of MID adoption within this sample; the two profiles are best understood as interpretive heuristics for communication and curriculum design rather than as discrete practitioner types.

### Network-based insights into MID-related constructs

Network analysis provided complementary insights into the interconnections among MID-related constructs. Although the estimated network was moderately sparse and centrality indices should be interpreted cautiously due to limited stability, several patterns remain informative for dental education. First, familiarity with MID emerged as the most interconnected construct, showing links with efficacy beliefs, caries classification, preventive knowledge, diagnostic knowledge, and MID-supportive attitudes. Although this pattern should not be interpreted causally, it suggests that familiarity with MID may provide an organizing point for connecting knowledge, beliefs, and practice-related dimensions. This underscores the value of embedding MID principles clearly and coherently in dental education and training.

Second, the MID-consistent *general practices* composite was directly connected to preventive knowledge, training exposure, and positive attitudes toward MID (dimensions that are plausible targets for educational intervention) as well as to the MID-aligned diagnostic methods composite. Finally, the strongest raw association in the network connected diagnostic knowledge with diagnostic methods used in practice. This finding, together with the low centrality of *diagnostic methods*, suggests that even when other MID-related dimensions are present, the connection between diagnostic knowledge and diagnostic methods identifies diagnostic reasoning as an important area for further educational research, although the present cross-sectional data cannot determine whether uncertainty about diagnostic tools constrains clinical decision-making.

From a curriculum design perspective, these conditional associations should be treated as hypotheses for future research rather than as evidence of causal intervention targets. They suggest that strengthening diagnostic knowledge and familiarity with MID principles could be examined within integrated training models, alongside outcomes in related knowledge, belief, and practice indicators. More broadly, these results align with international evidence indicating that although awareness of MID concepts is widespread, their translation into routine clinical practice remains uneven [6, 9, 12, 15, 24]. Future educational interventions should therefore evaluate training models that include repeated exposure to diagnostic reasoning, contemporary classification systems, and practical demonstrations, while longitudinal designs are needed to determine whether changes in these constructs produce broader effects across MID uptake. The role of mentoring and visible role modeling in challenging long-standing clinical habits should also be examined directly in future educational research.

### Strengths, limitations, and future directions

This study contributes to the literature on MID uptake in several ways. First, it introduces a profile-centered analytic framework that captures heterogeneity in MID uptake, offering a more nuanced alternative to mean-level, feature-by-feature comparisons. Second, integrating network analysis provided a complementary descriptive perspective on how MID-related constructs were conditionally interrelated. More broadly, the present study demonstrates how profile-centered and network-analytic approaches can be used to summarize multidimensional survey data on MID uptake and identify descriptive patterns for future research. This contribution is methodological and descriptive rather than explanatory: the analyses map how MID-related indicators co-occur and relate conditionally in this sample, but they do not identify causal mechanisms, barriers, or determinants of MID adoption. Although limited centrality stability cautions against treating specific nodes as established leverage points, the network findings can help generate more focused hypotheses about which components of MID uptake may warrant closer attention in future research. By linking these descriptive patterns to dental education and continuing professional development, the study aligns with current efforts to integrate MID principles more coherently into training contexts, while also pointing to questions that require future curriculum, implementation, or mixed-methods research.

Specifically, for educators and curriculum designers, these findings suggest that comprehensive MID training should move beyond isolated knowledge transfer and instead foster coherent competence systems that integrate diagnostic reasoning, clinical decision-making, and confidence in non-invasive and minimally invasive treatments. Embedding such training throughout dental education—rather than confining it to isolated modules—represents a plausible strategy for normalizing MID practices, but its effects on sustained adoption require direct evaluation. For researchers, replicating the present cluster and network analyses could be a useful approach to strengthen the international literature on MID knowledge, attitudes, and practices, with relevance to dental education.

Several limitations should be acknowledged. First, the cross-sectional design precludes causal inference regarding the directionality of associations among MID-related constructs. Longitudinal studies are therefore needed to examine how educational exposure and clinical experience shape trajectories of MID uptake over time. Second, the use of convenience sampling and self-reported measures may limit generalizability and introduce response biases. Although participants were recruited from multiple university centers and professional stages across Romania, future research would benefit from more representative sampling strategies and the inclusion of objective indicators of clinical behavior. Likewise, the final dataset did not retain respondent-level recruitment-source or university-center indicators, so when recruitment-period sensitivity analyses were conducted, timing effects could not be separated from unmeasured differences in recruitment context or sample composition.

Third, while the cluster solution was stable, the identified profiles were only moderately differentiated, and network centrality estimates showed limited stability. Accordingly, conclusions regarding discrete uptake profiles and the relative importance of specific constructs should be interpreted as exploratory. Replication with larger samples and alternative analytic approaches (e.g., latent profile analysis or different network estimation methods) would help strengthen confidence in these patterns.

Finally, although the survey instrument was developed through iterative expert review and the Romanian version was refined for conceptual clarity, the translation was not subjected to formal forward–back translation or independent linguistic validation, and the MID constructs were operationalized as formative indices rather than formally psychometrically validated scales. Likewise, alternative codings of “Not sure” responses were not examined; future validation work should test whether different coding schemes affect composite scores or profile/network results. Moreover, the emphasis on breadth across multiple MID domains necessarily limited the depth with which specific techniques, decision processes, or attitudinal nuances could be assessed. The survey also did not directly ask participants to identify perceived benefits of, or barriers to, MID implementation; therefore, the present analyses cannot explain individual motivations for embracing or resisting MID. Future studies could complement survey-based approaches with psychometrically validated measures, observational or performance-based assessments, and mixed-methods designs to further refine and validate the assessment of MID uptake in dental education and clinical practice.

## Conclusions

Despite broad professional endorsement of MID, its uptake in routine practice remains uneven and multifaceted. By combining a profile-centered clustering approach with network analysis, this study suggests that MID adoption can be usefully examined not in terms of the presence or absence of isolated skills, but as a configuration of interrelated competencies. The two moderately differentiated uptake profiles suggest that, in this sample, MID integration may be better understood along a continuum than as a set of sharply separated practitioner types, supporting inclusive rather than narrowly targeted educational strategies.

From an educational and implementation perspective, the findings support the relevance of embedding MID principles coherently across dental curricula and continuing professional development, with particular attention to diagnostic reasoning, contemporary classification systems, and confidence in preventive and minimally invasive approaches. Strengthening these elements as part of an integrated competence system—supported by hands-on training, mentoring, and visible role modeling—may offer a coherent direction for future curriculum development, but its impact on clinical practice should be tested in longitudinal or intervention studies.

## Data Availability

The de-identified analysis dataset and accompanying codebook are available on the Open Science Framework: https://doi.org/10.17605/OSF.IO/EP94Y

## Acknowledgments

The authors thank the PaFein+ project consortium for providing the broader educational context that facilitated this research, and the dental students, residents, and specialists who completed the survey.

## Supporting information

**S1 Appendix. Supplementary material.** This file contains quality checks for the formative MID composite variables, full item wording and response coding, statistical software details, recruitment-period sensitivity analyses, and a sensitivity network analysis including perceived peer norms for hygiene instruction as a node. The appendix includes Tables S1–S5 and Figs S1–S2.

**S2 Checklist. STROBE checklist for cross-sectional studies.**

## Notes

### Competing Interest Statement

The authors have declared no competing interest.

### Author Declarations

The study was conducted in accordance with the World Medical Association Declaration of Helsinki. This study was approved by the Ethical Commission of the Carol Davila University of Medicine and Pharmacy, Bucharest (Protocol No. 11580/30.04.2024). All participants received information about the study before completing the survey, and consent was provided by continuing to the next page and submitting the survey.

## References

1. International Association of Paediatric Dentistry. Minimal intervention dentistry: IAPD foundational articles and consensus recommendations. Available from: https://iapdworld.org/wp-content/uploads/2022/08/2022_02_minimal-invasive-dentistry.pdf. Accessed 2026 Jun 18.

2. BaniHani A, Santamaría RM, Hu S, Maden M, Albadri S. Minimal intervention dentistry for managing carious lesions into dentine in primary teeth: an umbrella review. Eur Arch Paediatr Dent. 2022;23(5):667–693. doi:10.1007/s40368-021-00675-6

3. Li X, Li R, Wang H, Yang Z, Liu Y, Li X, Xue X, Sun S, Wu L. Global burden of dental caries from 1990 to 2021 and future projections. Int Dent J. 2025;75:100904. doi:10.1016/j.identj.2025.100904

4. Tsakos G, Watt RG, Guarnizo-Herreño CC. Reflections on oral health inequalities: Theories, pathways and next steps for research priorities. Community Dent Oral Epidemiol. 2023;51:17–27. doi:10.1111/cdoe.12830

5. Abdelhafeez MM, Alharbi FM, Srivastava S, Eldwakhly E, Saadaldin SA, Soliman M. Perception of Minimum Interventional Dentistry among Dental Undergraduate Students and Interns. Medicina (Kaunas). 2023;59(4):649. doi:10.3390/medicina59040649

6. Dixit A, Sindi AS, Paul S, Badiyani BK, Kumar A, Arya R, Arora NN, Obulareddy VT. A Study to Assess Knowledge, Attitude, and Perception of Dental Practitioners on Minimally Invasive Dentistry Concepts. J Pharm Bioallied Sci. 2023;15(Suppl 2):S993–S996. doi:10.4103/jpbs.jpbs_255_23

7. Alyahya Y. A narrative review of minimally invasive techniques in restorative dentistry. Saudi Dent J. 2024;36(2):228–233. doi:10.1016/j.sdentj.2023.11.005

8. Perrone BR, Bottesini VC, Duarte DA. Minimal intervention dentistry: What is its clinical application and effectiveness in different continents? – A scoping review. J Conserv Dent Endod. 2024;27(2):134–139. doi:10.4103/JCDE.JCDE_274_23

9. Hafiz Z, Alhomaidhi M, Almutairi R, Alharbi A, Alshahrani L, Alzahrani S. Knowledge and attitudes of dental students and interns on minimally invasive dentistry for pediatric dentistry at King Saud University. BMC Oral Health. 2025;25(1):721. doi:10.1186/s12903-025-06087-y

10. Kumar S, Mala N, Rana KS, Namazi N, Rela R, Kumar K. Cognizance and Use of Minimally Invasive Dentistry Approach by General Dentists: An Overlooked Companion. J Pharm Bioallied Sci. 2021;13(Suppl 1):S199–S202. doi:10.4103/jpbs.JPBS_674_20

11. Global strategy and action plan on oral health 2023–2030. Geneva: World Health Organization; 2024. Licence: CC BY-NC-SA 3.0 IGO. Available from: https://www.who.int/publications/i/item/9789240090538. Accessed 2026 Jun 18.

12. de Moura RC, Santos PS, Matias PMDS, Vitali FC, Hilgert LA, Cardoso M, Massignan C. Knowledge, attitudes, and practice of dentists on Minimal Intervention Dentistry: A systematic review and meta-analysis. J Dent. 2023;132:104484. doi:10.1016/j.jdent.2023.104484

13. Mirsiaghi F, Leung A, Fine P, Blizard R, Louca C. An investigation of general dental practitioners’ understanding and perceptions of minimally invasive dentistry. Br Dent J. 2018;225(5):420–426. doi:10.1038/sj.bdj.2018.744

14. BaniHani A, Hamid A, Van Eeckhoven J, Gizani S, Albadri S. Minimal intervention dentistry (MID): Mainstream or unconventional option? Exploring the impact of COVID-19 on paediatric dentists’ views and practices of MID for managing carious primary teeth in children across the United Kingdom and European Union. Eur Arch Paediatr Dent. 2022;23:835–844. doi:10.1007/s40368-022-00746-2

15. Elsenberg CPM, van der Veen MH, Kwakkenbos L, Vreeken DD, Lagerweij MD, Volgenant CMC. Knowledge and attitude regarding preventive and minimally invasive caries treatments: A cross-sectional survey among Dutch dental professionals. Eur Arch Paediatr Dent. 2025;26:1–11. doi:10.1007/s40368-025-01130-6

16. Borsboom D, Deserno MK, Rhemtulla M, Epskamp S, Fried EI, McNally RJ, Robinaugh DJ, Perugini M, Dalege J, Costantini G, Isvoranu AM. Network analysis of multivariate data in psychological science. Nat Rev Methods Primers. 2021;1:58. doi:10.1038/s43586-021-00055-w

17. Hartigan JA, Wong MA. Algorithm AS 136: A K-means clustering algorithm. J R Stat Soc Ser C Appl Stat. 1979;28(1):100–108. doi:10.2307/2346830

18. Benjamini Y, Hochberg Y. Controlling the false discovery rate: a practical and powerful approach to multiple testing. J R Stat Soc Series B Stat Methodol. 1995;57(1):289–300.

19. Blanken TF, Isvoranu AM, Epskamp S. Estimating network structures using model selection. In: Isvoranu AM, Epskamp S, Waldorp LJ, Borsboom D, editors. Network psychometrics with R: A guide for behavioral and social scientists. 1st ed. Abingdon (UK): Routledge; 2022. p. 111–132.

20. Friedman J, Hastie T, Tibshirani R. Sparse inverse covariance estimation with the graphical lasso. Biostatistics. 2008;9(3):432–441. doi:10.1093/biostatistics/kxm045

21. Isvoranu AM, Epskamp S. Which estimation method to choose in network psychometrics? Deriving guidelines for applied researchers. Psychol Methods. 2023;28(4):925–946. doi:10.1037/met0000439

22. Epskamp S, Borsboom D, Fried EI. Estimating psychological networks and their accuracy: A tutorial paper. Behav Res Methods. 2018;50:195–212. doi:10.3758/s13428-017-0862-1

23. Fontana M, Gonzalez-Cabezas C, Tenuta LMA. Evidence-based approaches and considerations for nonrestorative treatments within modern caries management: Integrating science into practice. J Am Dent Assoc. 2024;155(12):1000–1011. doi:10.1016/j.adaj.2024.09.007

24. da Mata C, McKenna G, Hayes M. Knowledge transfer on the use of atraumatic restorative treatment: A mixed-methods study. J Dent. 2022;118:103944. doi:10.1016/j.jdent.2022.103944

